# Managing very small scale objective structured clinical examinations - can we ensure reliability of pass/fail decisions with low candidate numbers? A feasibility study of global score only decision making

**DOI:** 10.1101/2023.03.21.23287534

**Authors:** James Gray, Claire Darling-Pomranz, James Rone, David Spencer

**Author notes:** Corresponding author: James Gray, Academic Unit of Medical Education, The Medical School, The University of Sheffield, Beech Hill Road, Sheffield, South Yorkshire, S10 2RX, United Kingdom. Ethics approval Ethical approval was granted by the University of Sheffield ethics review committee on the 30^th^ of January 2020. University reference 032488.

## Abstract

**Background:** This study assesses a change to pass/fail decision making from an Angoff method to one based on examiner global judgements in very small scale Objective Structured Clinical Examinations (OSCE).

**Approach:** Data was collected from a Physician Associate OSCE in which two examiners marked each station one utilising a global rating based mark scheme and the other the original, Angoff based, mark scheme. Alternative global ratings were used to try and improve the judgement decision of examiners and checklists significantly reduced into a small number of broader domains. Examiners were also asked to provide some feedback on the new rating scales.

**Evaluation:** Mann-Whitney U tests were used to evaluate the data. Overall there was fairly good correlation of candidate performance between the two methods. Some anomalies were found with the use of the word safe in the global ratings creating a “killer” station. Examiners had mixed views but most were positive about the change in approach.

**Implications:** The initial results seem promising suggesting that global rating scales alone may be suitable to determine pass/fail decisions in very small scale OSCE with implications for all educators managing such assessments. We intend to keep the adjusted global rating scales.

## 1. Background

Standard setting of OSCE examinations has multiple approaches but in large number cohorts borderline regression is commonly utilised ensuring that the assessor/student interaction that has primacy in line with best practice (1). The University of Sheffield Physician Associate (PA) course is a very small cohort of 20 to 22 students thus the borderline regression methodology becomes statistically unsound. Studies looking at the use of borderline regression in small-scale OSCEs are in cohorts of around 50 to 60 students, much larger than our group (2-4). Angoff methodology is currently utilised with expert assessment of the OSCE station to determine an appropriate pass mark prior to the examination (5). It is notable that Regehr et al found that global judgement from expert assessors was the most reliable component of each OSCE station and contributed more to the overall reliability a checklist thus challenging this approach (6).

If focusing on global rating it seems logical that we should align the judgement sought from assessors to the question we would ask in “real world” scenarios which are seldom pass or fail. A large multicentre trial demonstrated an improvement in assessor decision making in workplace based assessment when this alignment was in place and others have commented on its potential to improve global rating scales in OSCEs (7).

This study aimed to evaluate the feasibility of using a Global Rating approach for pass/fail decisions in very small scale OSCEs.

## 2. Approach

The study was conducted in a formative Objective Sructured Clinical Examination (OSCE) run under full examination conditions in March 2020. The OSCE utilised 10 stations that had pre-agreed Angoff scores to ensure consistency with previous examinations.. The participants were a single cohort (n=19) of final year Physician Associate students from the University of Sheffield (UK). The sample size was small as this was a feasibility study specifically targeting very small scale OSCEs.

The examiners were from our trained examiner pool. The students were assessed by two examiners concurrently with one using the traditional mark scheme and one the new mark scheme. Two examiners were recruited for each station with one utilising the original, checklist focused, mark sheet whilst the second utilised a new mark sheet focussed on a small number of domains e.g. information gathering but with the global rating given primacy.

The measured variables were the student scores and global rating scales.To enable some numerical comparison the marks originally attributable to checklist areas under each domain were grouped to permit comparison of scores between the two. The original global rating scale had 5 points (Outright Fail, Borderline, Pass, Good Pass, Excellent Pass). In the new mark scheme the global rating anchors were changed to terms in line with the decision being sought (Cannot undertake this skill at a safe level, Can undertake this skill safely and Undertaking this skill at a high level) with no “borderline” option.

To improve validity and reduce bias the examiners were unaware of the Angoff cut scores and the mark sheets did not show the scoring schedule. Prior to the exam commencing examiners were permitted to discuss between themselves and “calibrate” their views on what constituted a safe or passing student. Once the exam was in progress examiners were not permitted to discuss their marking.

## 3. Evaluation

Initial results were analysed via direct comparison of pass/fail decisions between the two groups. In order to investigate the question as to what happens when the examiners have to make a pass/fail decision we investigated what we called the “marginal group”. This is a group of candidates who just achieved the Angoff score for a station or were either one or two marks below the score. The rationale is that a fail under the Angoff method is determined by the score rather than the global rating so we might consider the marginal group to be those that are close to the pass mark. The assigned numerical values of the global ratings (adjusted for the new method as per table 1) were used to compare the mean global rating scale (GRS) via the Mann-Whitney U test. Examiners using the new rating scales were also asked for feedback regarding the change of wording.

**Table 1.**
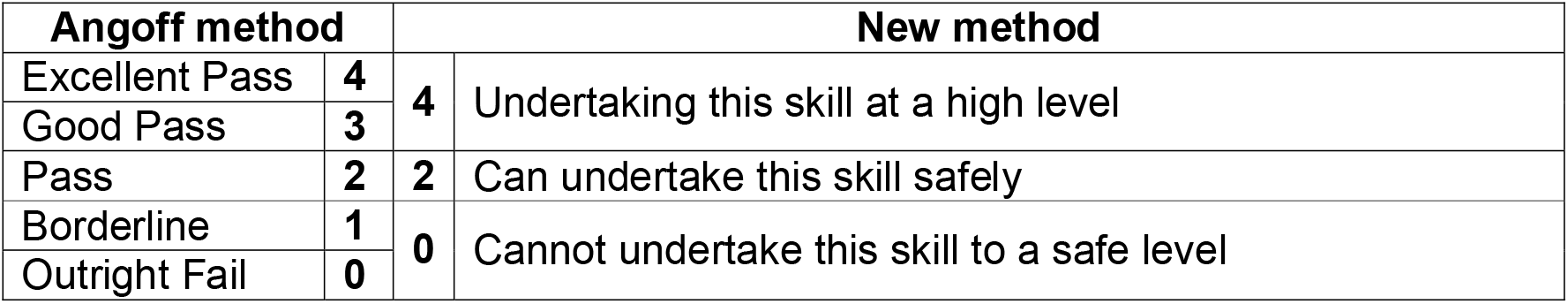
The assigned numerical values of the global ratings for statistical comparison

## 4. Results

### 4.1 Pass/Fail Results

During the study technical issues during station one led to some missing data from the new method and thus station one was omitted from the study. Table 2 shows the number of fails by station for each of the methods together with the number of fails in common between the two methods. Based on the results of both methods all students were considered to have passed the examination overall. The new method led to less station fails overall.

**Table 2:**
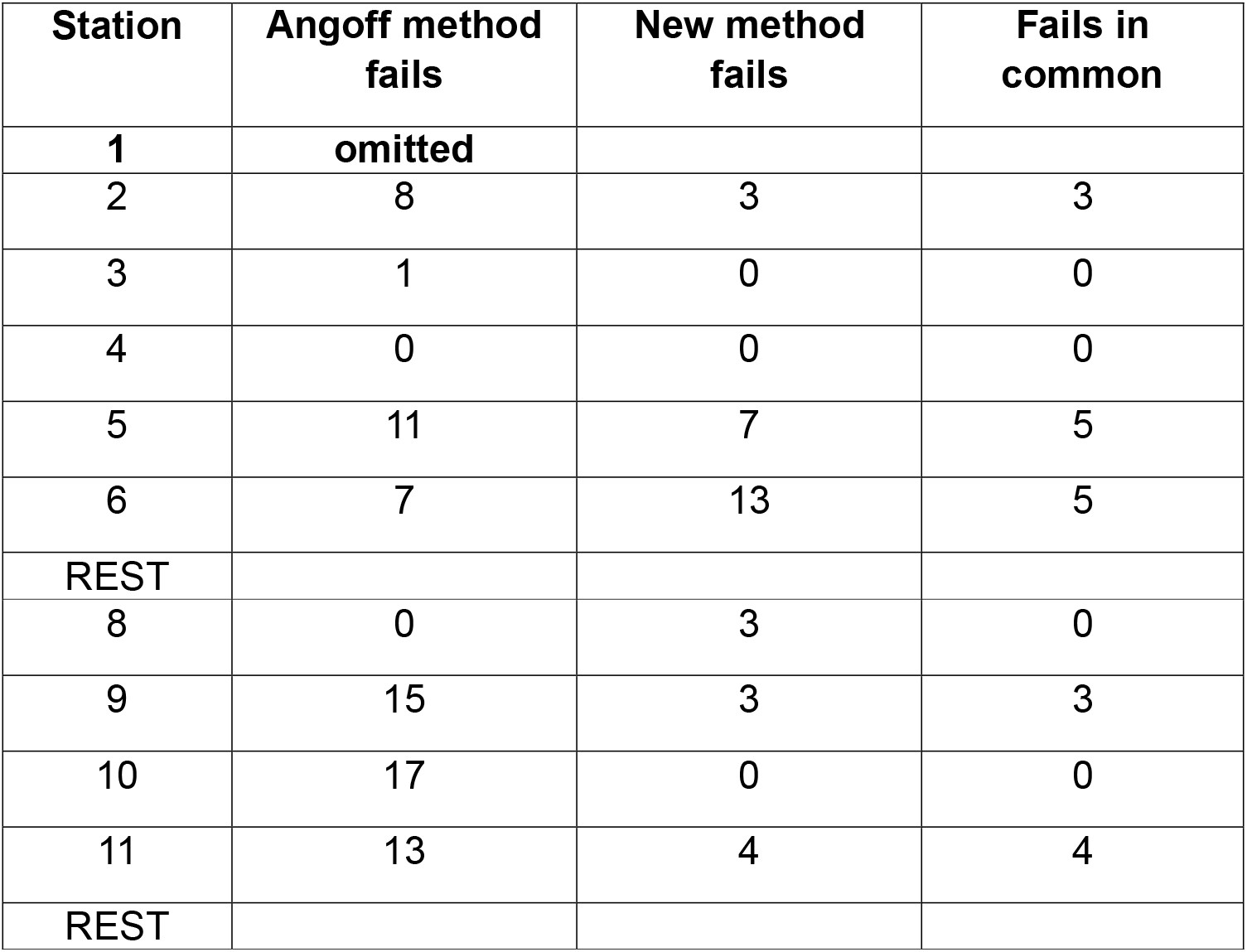
Number of fails for the OSCE stations under each method

**Table 3:** Examiner comments on the altered global rating scales

|  |  |
| --- | --- |
| **Benefits** | I felt empowered to make general judgements about the candidate |
|  | In the original system the student who asked every question they can think of without realising which are more relevant will often get points on the narrow banded sections like 'Enquires about Social history or FH [family history] or DH [drug history]' without showing that some of the answers have high relevancy |
|  | It is a more appropriate judgement of whether a PA is safe to start practicing |
|  | I think this is much more applicable to the overall question we're being asked to answer as examiners i.e. is this person safe to be let loose in clinical practice rather than the rather abstract question of whether they can perform well in an OSCE or not |
| **Concerns** | With the new system there is no way of demonstrating that a student is borderline. Their performance may not pose a safety issue but it might not be quite up to the level that would be expected yet one would be loathed to call it an 'outright fail' |
|  | I found it very uncomfortable to score someone as safe if e.g. they hadn't excluded red flags whereas I might have otherwise have recorded this as borderline or even pass if everything else ok (and one skill being assessed amongst several others) |
|  | I suspect the older marking scheme also acts as an aide memoire for examiners, easy to 'tick off' items as performed by |
|  | candidate and hence less likely to forget whether e.g. they asked re allergies |
|  | I liked this making scheme but it may need more training of examiners to do it on a larger scale |

In station ten, there were seventeen Angoff fails whereas the new method passed all students. Of those students failing under the Angoff method nine received a “Pass” global rating suggesting that the differences may have been due to examiner variability. There was generally an overlap of failing students i.e. the methods fail the same students, likely to be the weaker candidates.

The new method failed significantly more students in station six. The station six scenario involved a telephone call with a patient who may be having a myocardial infarction and should be advised to call an ambulance. Several students covered most items on the Angoff checklist, therefore passing the station under the Angoff method, however very few advised the patient to call an ambulance. Because of this, the examiners using the new method considered this “unsafe” and thus many students failed. This raises the question of whether the wording “safe” in the global ratings leads to potential “killer” stations.

Of the forty-five station scores included in the marginal group thirty-two were awarded a passing global score (safe or higher) under the new method whilst thirteen were not (roughly 2.5:1). These figures compare to roughly 1:1 passing vs failing for final year medical students given a borderline global rating in their OSCEs. The mean global rating scale (GRS) score was evaluated using the assigned numerical values of the global ratings under the two methods. This showed no difference in the mean GRS in all but one station. In general the correlation was fairly strong, however there were a few cases where there was no correlation, expected due to the scales both being small and the new GRS only having 3 points (0,2,4).

### 4.2 Examiner views on new rating scales

We asked examiners using the new mark scheme their opinions on the new rating schemes with eight responses received. Seven of them felt the new mark scheme allowed the student performance to be considered holistically more easily and potentially reward those who clearly are good at the process without necessarily ticking every box. It was noted that whilst this was easy for those clearly high, or poorly, performing it was more difficult with candidates typically perceived as “borderline” and the need to now make a pass/fail decision.

Six of the eight assessors felt that the new global rating terms were more appropriate than pass/fail etc. although the issue of the use of the word safe was brought up as a potential issue as noted in the overall scores.

## 5. Implications

Overall there was fairly good correlation of candidate performance between the two methods suggesting that using global ratings alone may be feasible for pass/fail decisions in very small scale OSCE.

Most of the unusual results seem to be due to either high Angoff pass marks or examiner variability. The issue of the Angoff method is highlighted with stations nine and eleven showing most students failing under the Angoff method but receiving a pass for their global rating. It has been noted that the Angoff method has tended to give high pass marks in written tests. Experts, as required by Angoff, tend to be more stringent thus the pre-test cut score is influenced by the make-up of the panel (8). If we can assume this applies equally to the OSCE it might explain the variance between the Angoff and GRS results, this emphasises the views that Angoff may be rewarding the ability to deliver against a checklist rather than a holistic, high quality, clinical performance. We believe that this has important implications for institutions still utilising the Angoff method to standard set OSCE stations as it would suggest that it is not the behaviours that we wish to see that are being rewarded and consideration should be given to discontinuing this approach to standard setting OSCEs.

### 5.1 Limitations / Generalizability

This feasibility study is limited by the small sample size and the challenge of statistical evaluation however the results should be generalizable to other very small scale OSCEs examinations. Based on the results all students were considered to have passed the examination under both methods. It is useful to note that candidates who undertook our OSCE went on to sit a national examination run by the Royal College of Physicians standard set using the borderline regression method. This cohort’s results showed only a single failure at the national examination. If our method was problematic and “over-scoring” we would have expected a higher rate of fails. This suggests that our standard is appropriately set by the examiners.

### 5.2 Limitations / Generalizability

Due to the small sample size further work needs to be carried out before we can be fully confident that the approach of using the global rating scales to make pass/fail judgements is fully secure but the initial results suggest that this is feasible. We intend to keep the adjusted global rating scales, albeit modified, and remove borderline permanently in these small scale examinations.

## Data Availability

All data produced in the present study are available upon reasonable request to the authors

## Notes

Conflict of Interest The authors have no conflict of interest to declare

### Competing Interest Statement

The authors have declared no competing interest.

### Funding Statement

This study did not receive any funding

### Author Declarations

Ethics Committee of The University of Sheffield (UK) gave ethical approval for this work. Ref 032488

